# Using 2D Video-based Pose Estimation for Automated Prediction of Autism Spectrum Disorders in Preschoolers

**DOI:** 10.1101/2021.04.01.21254463

**Authors:** Nada Kojovic, Shreyasvi Natraj, Sharada Prasanna Mohanty, Thomas Maillart, Marie Schaer

## Abstract

Clinical research in autism has recently witnessed promising digital phenotyping results, mainly focused on single feature extraction, such as gaze, head turn on name-calling or visual tracking of the moving object. The main drawback of these studies is the focus on relatively isolated behaviors elicited by largely controlled prompts. We recognize that while the diagnosis process understands the indexing of the specific behaviors, ASD also comes with broad impairments that often transcend single behavioral acts. For instance, the atypical nonverbal behaviors manifest through global patterns of atypical postures and movements, fewer gestures used and often decoupled from visual contact, facial affect, speech. Here, we tested the hypothesis that a deep neural network trained on the non-verbal aspects of social interaction can effectively differentiate between children with ASD and their typically developing peers. Our model achieves an accuracy of 80.9% (F1 score: 0.818; precision: 0.784; recall: 0.854) with the prediction probability positively correlated to the overall level of symptoms of autism in social affect and repetitive and restricted behaviors domain. Provided the non-invasive and affordable nature of computer vision, our approach carries reasonable promises that a reliable machine-learning-based ASD screening may become a reality not too far in the future.

## Introduction

Autism spectrum disorders (ASD) are a group of lifelong neurodevelopmental disorders characterized by impairments in social communication and interactions, and the presence of restricted, repetitive patterns of interests and behaviors [1]. Despite advances in understanding the neurobiological correlates of these disorders, there is currently no reliable biomarker for autism, and the diagnosis uniquely relies on the identification of behavioral symptoms. Although ASD can be detected as early as 14 months [2] and with high certitude before two years of age [3], the latest prevalence reports reveal that more than 70% of the affected children are not diagnosed before the age of 51 months [4]. Even in the absence of a highly specialized intervention program, earlier diagnosis is associated with a significantly better outcome. Indeed, specific strategies can be deployed to optimally support the child’s development during a period of enhanced brain plasticity [5]. Previous studies have demonstrated a linear relationship between age at diagnosis and cognitive gain [6, 7], whereby children diagnosed before the age of two years can gain up to 20 points in intellectual quotient (IQ) on average over the first year following diagnosis, while children diagnosed after the age of four will not show any substantial cognitive gain even with adequate intervention [7]. An efficient early screening, followed by early diagnosis, is the cornerstone to timely intervention. Most currently used screening tests are questionnaire-based, performing with low to moderate accuracy [8]. Further, they are prone to recall and subjectivity bias [9]. To overcome these limitations, tools that can deliver objective and scalable quantification of behavioral atypicalities are needed, particularly for the early detection of the signs indicative of autism.

A growing number of studies focus on the objective quantification of behavioral patterns relevant for autism, using the advances in machine learning (ML) and computer vision (CV) (for a review see [10]). For instance, Hashemi and colleagues developed an automatized CV approach measuring the two components of an early screening test for autism [12]. By tracking facial features, they automatically measured head turn to disengage attention from an object and head turn to track a moving object visually, the behaviors that previously were scored only manually. Another study using a name-calling protocol coupled with CV corroborated the well established clinical finding that toddlers with ASD respond less frequently when their name is called [13]. Additional to the automation of well established behavioral coding procedures, the use of these advanced technologies has allowed more subtle, dynamic measures of behavioral markers that would otherwise elude the standard human coding. Indeed, applying CV to the name-calling protocol revealed that, when children with ASD respond to their name, they tend to do so with an average of a one-second delay compared to the typically developing children [13]. In other studies, the use of motion capture and CV allowed to measure the reduced complexity of emotional expression in children with ASD, especially in the eye region [14]. Additionally, the combined use of motion capture and CV have provided insights on (i) the atypical midline postural control in autism [15, 16], (ii) highly variable gait patterns in ASD [17] and (iii) unique spatio-temporal dynamics of gestures in girls with ASD [18] that has not been highlighted in standard clinical assessments. Altogether, these studies demonstrate how computer vision and machine learning technologies can advance the understanding of autism, as they have the potential to provide precise characterizations of complex behavioral phenotypes.

The studies using ML and CV made a substantial contribution to the understanding of the disorder, offering a precise, objective measure of behavioral features that were traditionally assessed mostly qualitatively, if at all. However, there is still an important work to be done to enhance the scope and scalability of this approach. Most of the studies in this domain used fairly small samples, addressing rather specific questions focusing on one individual at time and measured behaviors elicited in controlled contexts [10]. A recent study undertook an effort to deploy a more holistic approach and, besides measuring the unique signature in the child’s behavior pattern, also focused on the child’s relation to immediate social context [19]. The authors used motion tracking to measure the approach and avoidance behaviors and the directedness of children’s facial affect during the diagnostic assessment - the Autism Diagnosis Observation Schedule (ADOS, [20, 21]). With these objective measures, the authors accounted for 30% of the standardized scores measuring the severity of autistic symptoms from only 5-minute excerpts of the free play interaction with the examiner. These results are auspicious as they do not focus on an individual in isolation but are a product of a more complex effort, the dynamic measure of the child’s relatedness to the social world. There is a critical need to take a more holistic stance to tackle the complex task of measuring how the child with autism interacts socially in settings close to everyday situations to advance towards a fully ecological and scalable approach.

Here, we present a machine learning algorithm to discriminate between ASD and typically developing (TD). From videos, acquired in our larger study on early development in autism, which feature social interactions between a child (with autism or TD) and an adult, we trained a deep neural network over the gold standard diagnostic assessment [20, 21]. The dimensionality of the input videos was reduced applying the multi-person 2D pose estimation OpenPose technology [22] to extract skeletal keypoints for all persons present in the video (see Fig. 1). Following [23], we then applied the CNN-LSTM architecture sensible to action recognition. Our goal was to explore the potential of purely non-verbal social interactions to inform automated diagnosis class attribution. The data included in this study comprised a Training set (34 TD children and 34 from children with ASD), and two validation samples, namely Testing Set 1 (34 from typically developping-TD children and 34 from children with ASD) and Testing Set 2 (*n* = 101, uniquely children with ASD) (see Table S1). The trained model distinguished children with ASD from TD children with an accuracy exceeding 80%. These results hold potential in accelerating and automatizing autism screening approach, in a manner that is robust and only minimally influenced by video recording conditions.

**Figure 1.**
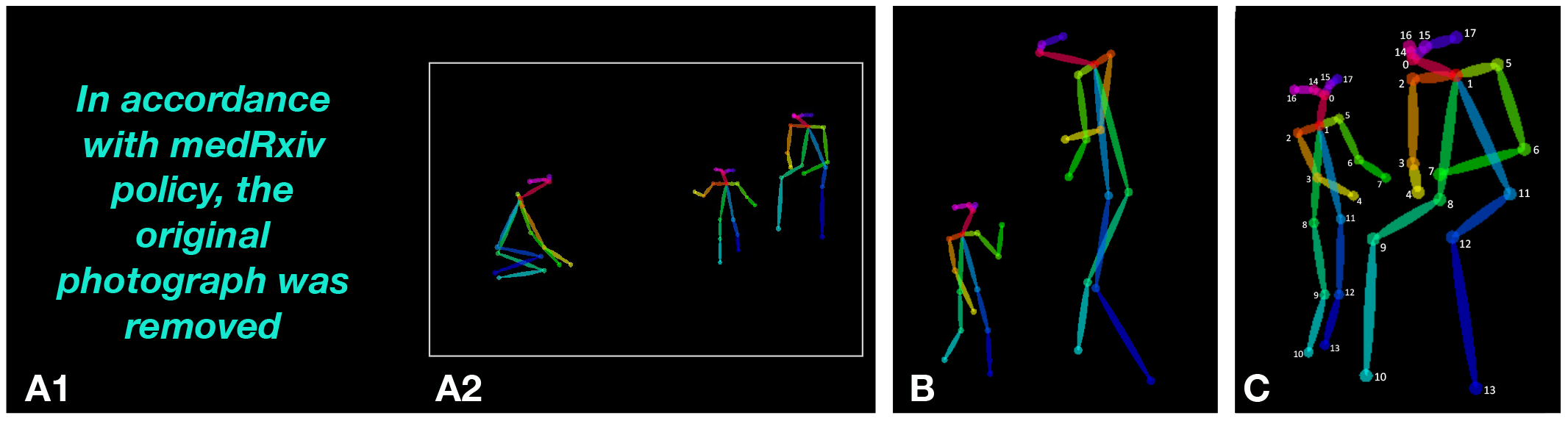
Example of 2D pose estimation using OpenPose on ADOS video frames: A1. OpenPose keypoints overlaid a video recording from the ADOS assessment, A2. OpenPose skeletal keypoints plotted over a null background, B. Example of *requesting* behavior with skeletal points, C. Example of *showing* behavior with numerated keypoints. Keypoint list: 0 = nose, 1 = heart, 2 = right shoulder, 3 = right elbow, 4 = right wrist, 5 = left shoulder, 6 = left elbow, 7 = left wrist, 8 = right hip, 9 = right knee, 10 = right ankle, 11 = left hip, 12 = left knee, 13 = left ankle, 14 = right eye, 15 = left eye, 16 = right ear, and 17 = left ear.

## Results

The final model architecture was obtained upon testing various configurations (see Fig.S2 and Supplementary section). The retained model was trained over the Training Set videos (68 ADOS videos, equally split between ASD and TD groups; see Table S1) that contained solely skeletal information on the black background, without sound (see Fig.1). Figure 2, Figure S1, the Methods and Supplementary sections detail different stages of the model training and validation. The predictions were obtained for individual 5s video segments and aggregated over the entire ADOS video for each subject from the two testing sets (see Fig.2). We further examined the stability of the diagnosis prediction as a function of the video input length. Finally, we explored the potential of a non-binary, continuous value of ASD probability to capture meaningful clinical characteristics, examining whether standardized scores obtained from various gold-standard clinical assessments related to the ASD probability extracted from the neural network.

**Figure 2.**
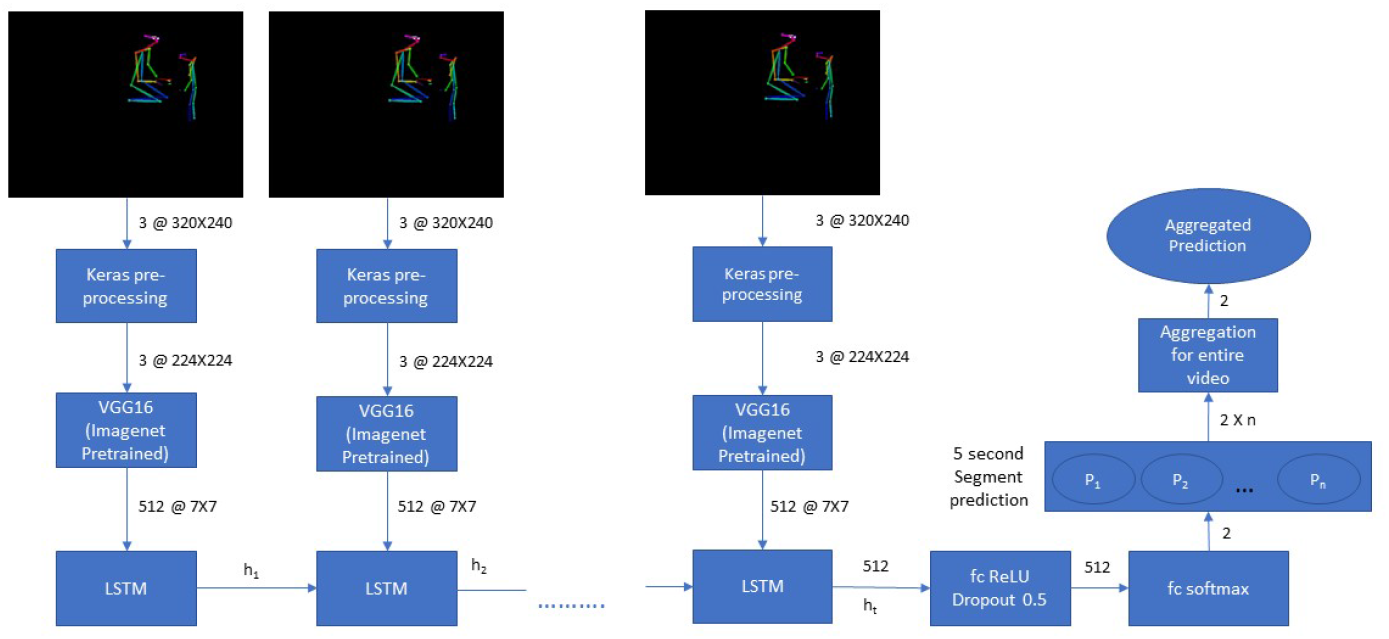
Neural network architecture. The pretrained Convolutional Network VGG16 [23] was used to extract the characteristics of all videos split into 5s segments. The output from this feature extraction step was fed into a LSTM network operating with 512 LSTM units. Finally, the output of the LSTM was followed by 512 fully connected ReLU activated layers and a softmax layer yielding two prediction outputs for the given segment. The segment-wise classifications were aggregated for the video’s duration to obtain a final prediction value (ranging from 0 to 1) that we denote “ASD probability”. The video was classified as belonging to a child with ASD if the mean value of ASD probability was superior to 0.5.

### Prediction of autism

Our model achieved an F1 score of 0.818 and a prediction accuracy of 80.9% over a sample of 68 videos in Testing Set 1 (Table 1). The same trained model achieved a prediction accuracy of 80.2% over a Testing Set 2 comprising 101 videos from children with ASD, thus endorsing the model’s stability.

**Table 1.** Accuracy, Precision, Recall, Specificity and F1 score for Testing set 1 predictions using VGG16 LSTM trained model at 80-20 training-validation split, 100 epochs, 128 batch size

| Parameter | Model (80-20 split) |
| --- | --- |
| Accuracy | 0.809 |
| Precision (Positive Predicted Values) | 0.784 |
| Recall (Sensitivity) | 0.854 |
| Specificity | 0.765 |
| F1 Score | 0.818 |

### Consistency of the ASD prediction over the video length

We further tested the extent to which the video length influenced our model’s prediction accuracy. By varying the length of the video input in the Testing set 1, we demonstrated that an average 70% accuracy is already obtained with 10 min video segments (see Fig.3A). As shown in Figure 3B, the prediction consistency is also very high across the video of a single individual, even with relatively short video segments. For instance, for half of the ASD sample, our method achieves a 100% consistency in prediction based on randomly selected 10 minutes segments. These results strongly advocate for the feasibility of video-based automated identification of autism symptoms. Moreover, the ADOS videos used in the present study were acquired using different systems. However, the accuracy of classification showed robustness to the variability in context, thus again highlighting the potential for generalization of this type of approach (see Supplementary section and Fig.S4).

**Figure 3.**
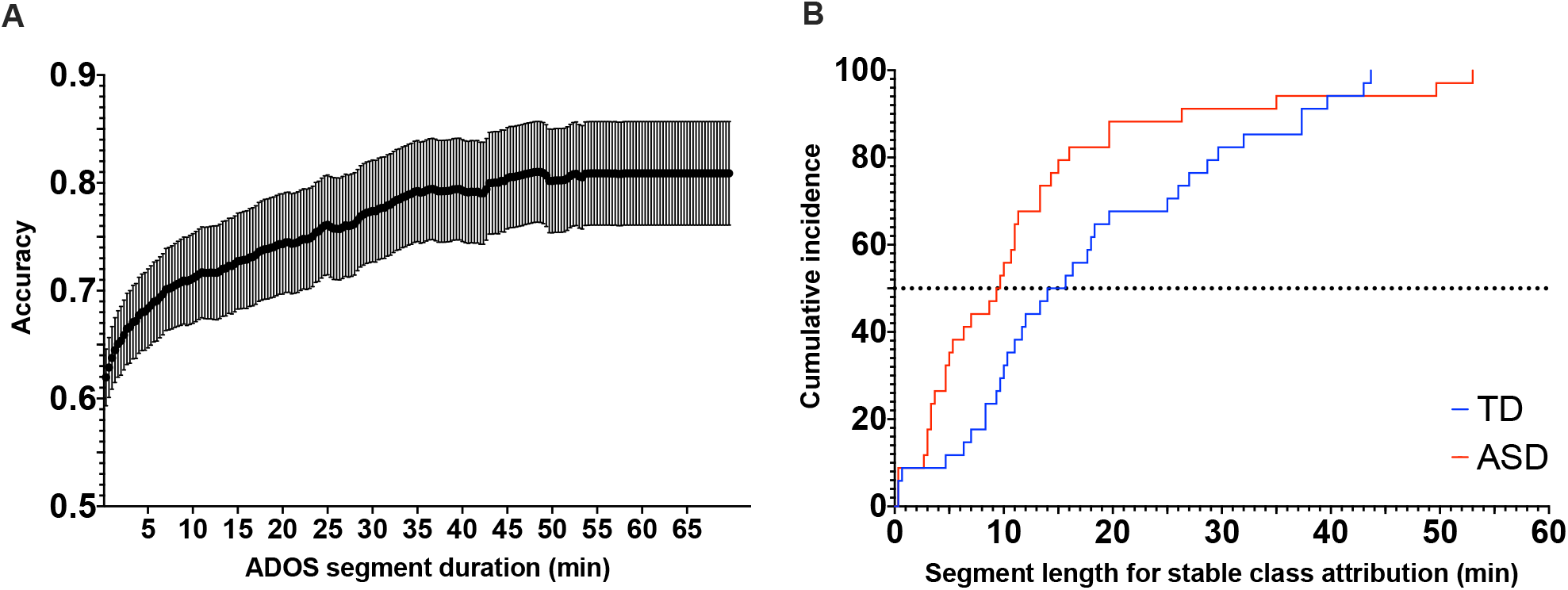
A. Association between the prediction accuracy and the length of considered video segment. The accuracy increases with longer video segments, with the final accuracy being 81% for Testing set 1 in our sample. B. Stability in the prediction as a function of the length of the considered video segment for the Testing set 1. The cumulative incidence depicts the required length of video segments that is needed to achieve 100% prediction consistency for all the segments of the same length randomly drawn from the full video of the same participant.

### Neural network derived ASD probability reflects the clinical phenotype

Using validated clinical assessments (Methods section), we then observed that neural network derived ASD ability was positively related to the overall level of symptoms of autism (*r*_s_(68) = 0.451, *p* < 0.001), (Fig. 4A), and this pattern was observed both in the domain of the social affect (*r*_s_(68) = 0.509, *p* < 0.001) and in the domain of repetitive and restricted behaviors (RRB) (*r*_s_(68) = 0.409, *p* < 0.001) (Fig.S5, panels A1-2). Moreover, ASD probability negatively correlated with the general adaptive functioning (*r*_s_(67) = −0.444, *p* < 0.001) (Fig.4,B). Further analyses revealed that ASD probability was related to the communication (*r*_s_(68) = −0.386, *p* < 0.001), socialization (*r*_s_(68) = −0.477, *p* < 0.001) as well as the autonomy in daily life (*r*_s_(68) = −0.397, *p* < 0.001) but not with the functioning in the motor domain (*r*_s_(68) = −0.186, *p* = 0.066) (Fig.S5, panels B1-3). Finally, ASD probability showed a moderate negative correlation with cognitive functioning (*r*_s_(63) = −0.283, *p* = 0.012) (Fig.4C).

**Figure 4.**
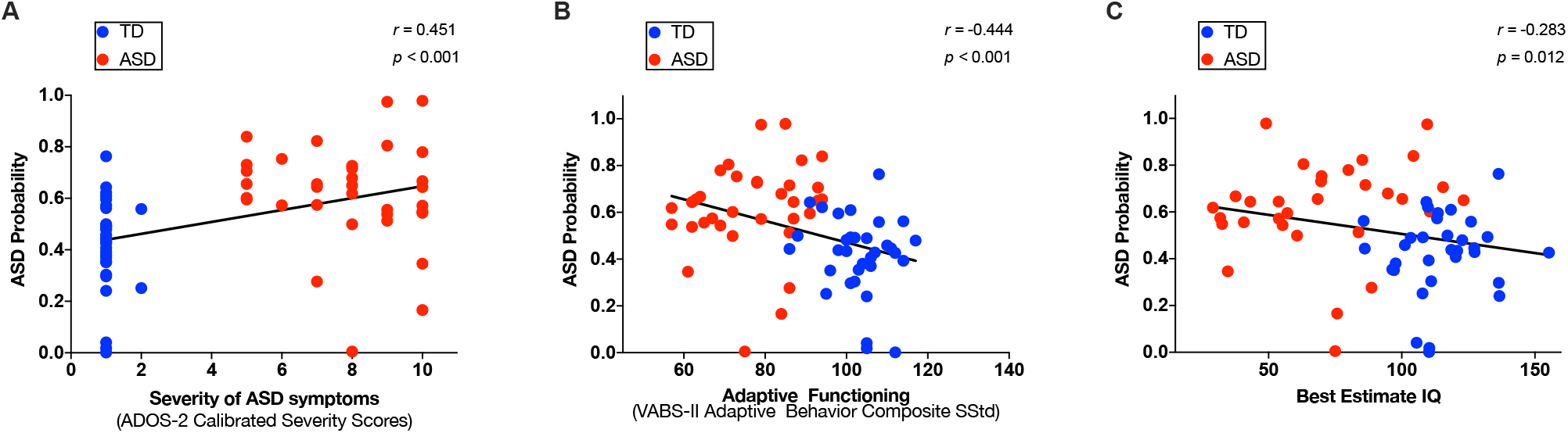
Scatter plots depicting the relation between predicted ASD probability and clinical measures: A. Total level of severity of autistic symptoms, B. Adaptive functioning, C. Best estimate Intellectual Quotient. The least squares linear fit is depicted as black line and values of Spearman r coefficient and corresponding p values are shown on each panel. Ground-truth classes: ASD=red, TD=blue.

The above correlations were based on ADOS severity scores, representing a summarized measure of the degree of autistic symptoms. In addition, we were interested in understanding how the automatically derived ASD probability related to individual autistic symptoms and potentially inform us about the symptoms that were more closely related to ASD class attribution. After applying Bonferroni corrections, ASD probability was positively related with three symptoms in the communication domain of ADOS, namely gestures (*r*_s_(68) = 0.435, *p* < 0.001), pointing (*r*_s_(68) = 0.540, *p* < 0.001) and intonation (*r*_s_(68) = 0.426, *p* < 0.001) (Fig.S6, panels A-C). In the social interaction domain of the ADOS the ASD probability was related to unusual eye contact (*r*_s_(68) = 0.500, *p* < 0.001), directed facial expressions (*r*_s_(68) = 0.488, *p* < 0.001), spontaneous initiation of joint attention (*r*_s_(68) = 0.450, *p* < 0.001), integration of gaze and other behaviors (*r*_s_(68) = 0.591, *p* < 0.001), giving (*r*_s_(68) = 0.438, *p* < 0.001), showing (*r*_s_(68) = 0.396, *p* < 0.001), shared enjoyment (*r*_s_(68) = 0.359, *p* = 0.001), quality of social overtures (*r*_s_(68) = 0.484, *p* < 0.001) (Fig.S6, panels D-K). Furthermore, ASD probability was positively related to functional play (*r*_s_(68) = 0.418, *p* < 0.001) and imagination (*r*_s_(68) = 0.470, *p* < 0.001)(Fig.S6, panels L-M). Finally, in the domain of repetitive behaviors and restricted interests, ASD probability was related to unusual sensory behaviors ((*r*_s_(68) = 0.434, *p* < 0.001) and unusually repetitive interests and stereotyped behaviors (*r*_s_(68) = 0.455, *p* < 0.001) (Fig.S6, panels N-O). The symptoms that were related to the ASD probability were predominantly non verbal. This finding strongly support the potential of our model to learn from the non verbal patterns of social interaction, clinically relevant in younger children.

## Discussion

Our neural network model operated on a low dimensional postural representation derived from social interaction videos between a child and an adult and robustly distinguished whether the child has autism, with a model prediction accuracy of 80.9% (precision: 0.784; recall: 0.854). We choose the model that operates on a relatively reduced set of characteristics, targeting non-verbal features of social interaction and communication, particularly relevant in very young children [20, 21]. We deployed an LSTM network learning temporal dependencies of relations between skeletal keypoints in 5s interaction segments to perform the classification. Our findings’ clinical validity was corroborated with positive correlations between neural network derived ASD probability and levels of autistic symptoms and negative correlations between the same measure and cognitive and everyday adaptive functioning of these children. Moreover, we showed that the accuracy of classification of around 70% was achieved based on only 10 minutes of filmed social interaction, opening avenues for developing scalable screening tools using smaller excerpts of videos.

The choice of the reduced dimensionality (2D estimated postural representations) in input videos was two-fold: to allow a pure focus on non-verbal interaction patterns and ensure de-identification of the persons involved (skeletal points plotted against the null background). To further promote the approach’s scalability, videos were not manually annotated; thus, we removed the human factor in the initial feature breakdown. Our design aligns with and expands findings from a smaller number of studies that probed automated video data classification [24, 25]. Zunino et al. [24] were able to determine a specific signature of grasping gesture in individuals with autism using the raw video inputs. This approach allowed a classification accuracy of 69% using 12 examples of grasping action per individual obtained in well-controlled filming conditions. In contrast, in our study, the input videos were remarkably heterogeneous in terms of recorded behaviors. They included moments of the interaction of a child with an examiner in the presence of caregiver(s). Moreover, different examiners performed the assessments as they are acquired as a part of the ongoing longitudinal study. Finally, regarding the pure physical aspects, these assessments took place in several different rooms and were filmed with different camera systems. Nevertheless, our study’s classification accuracy is superior to the one reported in the study using the controlled video of a very precise grasping action.

The clinical validity of our findings is supported by the significant correlation observed at the level of individual autistic symptoms (single items from the ADOS). The lack of ability to adequately integrate gaze to other communication means and impaired use of visual contact together with the reduced orientation of facial affect were the symptoms that were the most related to the neural network derived values of probability of ASD class attribution. Other symptoms that were strongly linked to the probability of receiving ASD class attribution comprise aberrant gesture use, unusual social overtures, repetitive patterns of behaviors and unusual sensory interests. These findings emphasize the potential of these low-dimensional social interaction videos to convey the atypicality of the non-verbal interaction patterns in young children with ASD. Indeed, out of 27 selected behaviors the 15 that were significantly related to the neural network derived ASD probability were the symptoms with a predominant non-verbal (e.g., giving, showing, spontaneous initiation of joint attention). This finding is in line with findings reported applying ADOS-like rating on home videos [26] who found that the aberrant use of visual contact was a feature that was the most determinant of ASD classification. Another study building an automated screener for autism symptoms based on annotated home videos reported that the screener capitalized on the non-verbal behaviors (e.g. eye contact, facial expressions) in younger participants while relying more on verbal features in older participants [27]. Indeed, clinically, the aberrant use of visual contact and aberrant gesture use are among the most striking and early emerging features of the disorder [28, 20, 21].

The major contribution of the automated identification of behaviors indicative of autism lies in enhancing the efficiency and sensitivity of the screening process. The diagnosis process is complex and delicate and is unlikely to be set on the track of automated performance before long. However, more informed, more objective and available screening is crucial to catalyze diagnosis referrals, hopefully leading to earlier intervention onset. Early interventions are of life-changing importance for individuals with autism. They improve their cognitive and adaptive functioning and reduce the severity of ASD symptoms [6, 29]. In the year following the diagnosis, children who receive early intervention – and start developing language skills before the age of 3 show the most important gains as young adults [30, 31].

Our results speak in favor of more objective, holistic, automatized methods as complementary tools to the ones used in clinical practice. In ASD, the availability of standardized measures of autistic symptoms was crucial in informing the clinic and the research [32]. Nevertheless, these gold-standard measures still rely on somewhat coarse descriptions of symptoms. Individual symptoms of autism are assessed on a 3 or 4 point scale [28, 20, 21] while phenotypical differences between the behaviors brought on the same plane can be very pronounced. The development and improvement in quantitative measures leading to a more fine-grained “digital phenotyping” [33] can be a tremendous asset in the early detection of signs of the disorder and the follow-up of its manifestation through development. Besides being more objective compared to human coding, it can allow the processing of larger quantities of the data and at the spatio-temporal resolution that is off limits to human coding. Moreover, these precise and continuous measures may uncover behavioral manifestations of the disorder that were previously not evidenced. They also may help define sub-types of the disorder to allow more precise clinical action [34]. The finding that we were able to achieve a robust accuracy of classification based on a limited set of characteristics derived from social interaction videos is very promising. This approach would further benefit from the implementation of the spatio-temporal attentional mechanism [35] to allow knowledge on the specific elements in space and time used to inform the diagnosis process in the network and improve our understanding of the manifestation of the disorder.

## Method

### Participants

The initial sample included sixty-eight children with autism (2.80 *±* 0.92 years) and 68 typically developing children (2.55 *±*0.97 years) who were equally distributed to compose the Training and Testing set (Testing Set 1), matched for diagnosis, age, gender and ADOS module (see Table S1). To validate the robustness of our classification method we included an additional testing sample comprising 101 videos from children with ASD (3.46*±*1.16 years) that we denote Testing set 2. All data used in this study were acquired as a part of a larger study on early development in autism. Detailed information about cohort recruitment was given elsewhere [36, 37, 38]. The current study and protocols were approved by the Ethics Committee of the Faculty of Medicine of the Geneva University, Switzerland. The methods used in the present study were performed in accordance with the relevant guidelines and regulations of the Geneva University. For all children included in this study, informed written consent was obtained from a parent and/or legal guardian. Children with ASD were included based on a clinical diagnosis according to DSM-5 criteria [1], and the diagnosis was further corroborated using the gold standard diagnostic assessments (see Clinical Measures subsection and Supplementary section). Typically developing (TD) children were screened for the presence of any known neurological or psychiatric illness and ASD in any first-degree relative of the child.

### Clinical measures

A direct measure of autistic symptoms was obtained using the Autism Diagnostic Observation Schedule-Generic ADOS-G, [20] or a more recent version Autism Diagnostic Observation Schedule-2^nd^ edition (ADOS-2) [21]. Cognitive functioning was assessed using various assessments depending on the children’s age and their ability to attend demanding cognitive tasks. We defined the Best Estimate Intellectual Quotient [39, 38] that combines the most representative cognitive functioning measures for each child. Adaptive functioning was assessed using the Vineland Adaptive Behavior Scales, second edition (VABS-II; [40]) (see Supplementary section for a detailed characterization of clinical measures).

### Video Data

To probe the diagnosis classification using machine learning on social interaction videos, we used filmed ADOS assessment acquired in the context of our study. Practical reasons drove this choice, ADOS being the most frequent video-based assessment in our study (systematically administered in all children included in our study). Moreover, ADOS provides a standardized and rich context to elicit and measure behaviors indicative of autism across broad developmental and age ranges [20]. Its latest version (ADOS-2) encompasses five modules covering the age from 12 months to adulthood and various language levels ranging from no expressive use of words to fluent complex language. To best fit the younger participants’ developmental needs, Modules Toddler 1 and 2 are conducted while moving around a room using a variety of attractive toys, while Modules 3 and 4 happen mostly at a table and involve more discussion with lesser use of objects. In this work, we focused uniquely on the Modules Toddler, 1 and 2, as these require fewer language abilities and are more sensitive to non-verbal aspects of social communication and interaction that we target using machine learning. The clinical findings on the prevalence of non verbal-symptoms in younger children [28, 20, 21] drove our choice to focus uniquely on non-verbal aspects of communication and interaction.

### Pose estimation

To purely focus on social interaction and essentially its non-verbal aspects, we extracted skeletal information on people present in ADOS videos using deep learning based multi-person 2D pose estimator-OpenPose [22]. OpenPose estimates keypoints of persons detected on the image/video independently for each frame. It uses a simple 2D camera input not requiring any external markers or sensors, thus allowing the retrospective analysis of videos. It also is immune to variations in resolutions and setting that images and videos might present. For the OpenPose output, we opted for the COCO model providing 2D coordinates of 18 keypoints (13 body and 5 facial keypoints; see Fig.1). The ordering of the keypoints is constant across persons and frames. Consistent with our focus on the non-verbal features of interaction during the semi-standardized ADOS assessment, we removed the background from all the videos and preserved only skeletal information for further analysis. To obtain feature vectors invariant to a rigid body and affine transformations and to increase the generalizability of our approach, we based our calculation on image output and not on raw keypoints coordinates (Fig.1) [41].

### Building the Neural Network

The OpenPose processed videos were down-sampled from 696 × 554 to 320 × 240 pixels and split into segments of 5s (see Table 2). To estimate the training and validation loss we used a categorical cross entropy loss function using a rmsprop optimizer. We found that the 5-second video segments were optimal for model training and resulted in less validation loss compared to longer segments (10s or 15s) (see Fig.S3). We opted for a CNN LSTM architecture for our model as it previously showed a good performance in video-based action classification ([42]). We used a VGG16 convolutional neural network ([43]), pretrained on the ImageNet ([44]) dataset to extract high dimensional features from individual frames of the 5 second video clips.The VGG16 is a 16 layers convolutional neural network that works with a 224×224 pixel 3 channel(RGB) input frame extracted from the video segment. The resolution is then decreased along the each convolution and pooling layer as 64 @ 112×112, 128 @ 56×56, 256 @ 28×28, 512 @ 14×14 and 512 @ 7×7 after the last convolution or pooling stage which has 512 feature maps. The high dimensional features extracted are flattened and input to a 512 hidden layered 0.5 dropout LSTM at a batch size of 128 ([45]) followed by fully connected dense layers with ReLU activation, 0.5 dropout and a softmax classification layer giving an 2 dimensional output (corresponding to the two classes, ASD and TD).

**Table 2.** Representation of video characteristics included in the Training and Testing set 1 as well as Testing set 2 used to validate the robustness of neural network derived classification

| Parameter | Training set | Testing set 1 | Testing set 2 |
| --- | --- | --- | --- |
| Number of Videos | 68 | 68 | 101 |
| Total duration (in hours) | 47.782 | 48.661 | 50.043 |
| Videos per Class | 34 | 34 | 101 (ASD) |
| Average Video Length | 42.16 min | 42.68 min | 41.965 min |
| Average number of 5 second segments per video | 505.92 | 512.16 | 503.574 |

The input training data of 68 ADOS videos were split in the ratio of 80-20, where the model used 80% of data for training and 20% of data was used for validation. We then analyze the model’s training and validation loss to avoid overfitting and perform hyperparameter tuning. The training and validation loss over a varied number of epochs is shown on Figure S3. The least validation loss model was deployed to predict over 5-second segment of the videos from the two testing sets (Testing Set 1 and Testing Set 2). We average the predictions for all the video segments to obtain a final prediction value denoted as “ASD probability”. We trained the neural network model over 5-second video segments at 128 batch size, 100 epochs and 80-20 training-validation split and used the trained model to make predictions over Testing Set 1 and Testing set 2 to check the accuracy of the prediction results across different testing sets.

The model training and validation was performed at University of Geneva High Performance Computing cluster, Baobab (Nvidia RTX 2080Ti, Nvidia Titan X and Nvidia Quadro P1000 GPUs).

### Statistical Analyses

The obtained measure of ASD probability derived from the neural network was further put in relation to standardized behavioral phenotype measures in children included in the Testing 1 sample. We calculated Spearman rank correlations with measures of severity of symptoms of autism (Total CSS, SA CSS and RRB CSS), adaptive (VABS-II Total and scores across four subdomains) and cognitive functioning (BEIQ) (Supplementary section). Furthermore, in order to obtain more fine-grained insight into the relation of ASD probability across the entire video and the specific symptoms of autism, this measure was correlated with raw scores on a selected set of 27 items that repeat across the three modules we used (for more details, please refer to Supplementary section). Results were considered significant at p < 0.05. The significance level was adjusted using Bonferroni correction for multiple comparisons. Thus the results concerning the two ADOS subdomains results were considered significant at p < 0.025 and four VABS-II subdomains at p < 0.013. For the analyses involving the 27 individual items of ADOS (for a full list please refer to Table S3), the results were considered significant at p < 0.002. For a comparison of the Training and Testing samples with regards to clinical assessments (ADOS, VABS-II and BEIQ), we used Student t-tests and Mann-Whitney tests when measures did not follow a normal distribution according to the D’agostino & Pearson test (See Table S1). Statistical analyses were conducted using GraphPad Prism v.8, (https://www.graphpad.com/scientific-software/prism/) and SPSS v.25 (https://www.ibm.com/analytics/spss-statistics-software).

### Relation of video length to prediction accuracy

Our final aim was to apprehend the length of video segments required for stable class attribution, thus probing the approach’s scalability. To this end, we employed a sliding window approach, starting with a length of 20 seconds and then stepwise increasing the window length by 20 seconds until window length matched video duration. In each window, ASD probability values are averaged over the containing 5-second segments for each of 68 videos in the Testing set 1 (Fig.3A). Using this method, we also test the prediction consistency for videos of a single individual class by identifying the sliding window length required for stable class attribution (Fig.3B).

## Data Availability

Upon publication in a peer-review journal, data can be obtained upon reasonable request

## Acknowledgments

We express our utmost gratitude to all families that took part in this study. We thank our colleagues for their precious contribution to data collection. Authors express a special gratitude to Ms Kenza Latréche who helped with manual annotation of recording settings.

Funding for this study was provided by National Centre of Competence in Research (NCCR) Synapsy, financed by the Swiss National Science Foundation (SNF, Grant Number: 51NF40_185897), by SNF grants to M.S. (#163859 and #190084), the UNIGE COINF2018 equipment grant, by the SDG Solution Space (https://www.fablabs.io/labs/sdgsolutionspace and by the Fondation Pôle Autisme (https://www.pole-autisme.ch).

## Supplementary Material

### Participants

The Table S1 summarizes the clinical characteristics of the Training and Testing 1 samples as well as additional Testing 2 sample that was used for validation of the model. Our initial sample of TD children included 68 children (28F). Then we selected the sample of 68 children with ASD to match the TD sample with regards to age and gender. Both samples (TD and children with ASD) were divided in two so that 34 children with ASD and 34 children with TD composed Training and Testing set respectively. Children with ASD included in both samples showed a moderate to a high level of autistic symptoms, as illustrated with their average ADOS-CSS of 7.47 and 7.88, respectively. No significant differences in all used clinical measures were found between Training and Testing 1 sample (see Table S1). The Testing 2 sample is composed of the videos that were available in our cohort but who were not entered in the initial sample due to the fewer number of available videos from TD children. The children in this sample were predominantly males (1F), presenting a moderate to severe levels of symptoms of autism (mean of 7.54) and mean cognitive scores of 74.8.

**Table S1.**
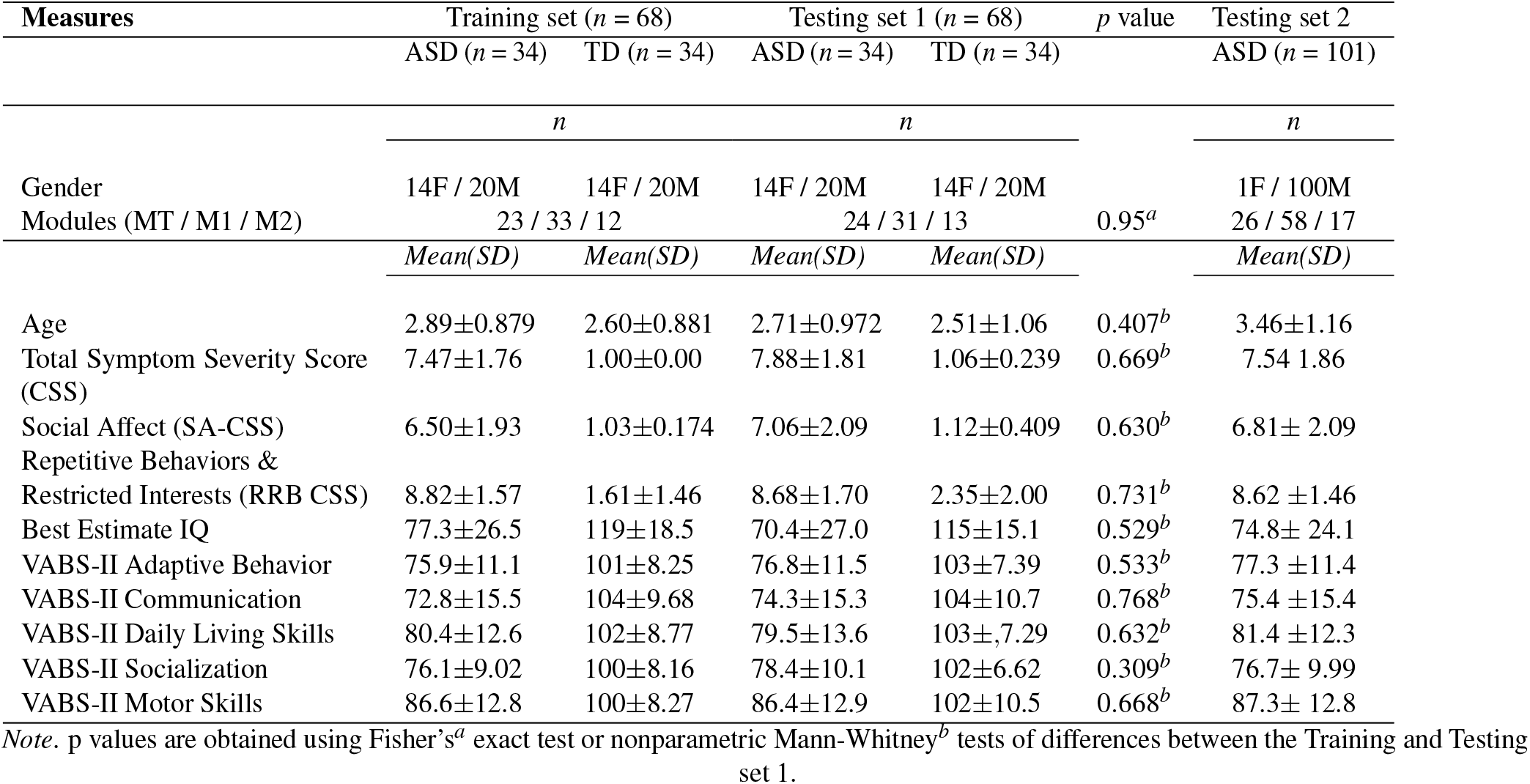
Description of the Training and two Testing sets of videos used in the present study.

### Clinical Measures/Behavioral Phenotype

A direct measure of autistic symptoms was obtained using Autism Diagnostic Observation Schedule-Generic ADOS-G, [20] or Autism Diagnostic Observation Schedule-2^nd^ edition (ADOS-2) [21]. Only one module is administered at one time. The latest version of ADOS allows to severity comparison scores ranging 1-10 that are aimed to be relatively independent on the participant’s characteristics such as age or verbal functioning [46]. For subjects who were administered the older version of the ADOS (ADOS-G), the scores were recoded according to the revised ADOS algorithm [47] to ensure comparability with the ADOS-2. Besides comparison severity scores that indicate the overall severity of autistic symptoms [46, 29] we also included the severity scores according to domains of social affect (SA) and restricted and repetitive behaviors (RRB) thus allowing for a more precise insight on the separate dimension of autistic symptoms [48].

Cognitive functioning was assessed using various assessments depending on the age of the children and their ability to attend demanding cognitive tasks. Inspired by previous work, we defined the Best Estimate Intellectual Quotient [39, 38]. For the majority of children Psycho-Educational Profile, third edition, Verbal/Preverbal Cognition scale (PEP-3; VPC DQ, [49]) (*n* = 49) was administered. For a smaller subset of children when PEP-3 was not administered we used Mullen Early Learning scales [50], (*n* = 14).

Adaptive functioning was assessed using the Vineland Adaptive Behavior Scales, second edition (VABS-II; [40]). This standardized parents interview measures adaptive functioning from childhood to adulthood in the domains of communication, daily-living skills, socialization, and motor domain. In each domain, standardized scores (SStd) are denoted by adaptive level, ranging from low to high. The four domain standardized scores are then combined to yield the adaptive behavior composite score as a global measure of adaptive functioning of an individual.

### Neural Network Training

The neural network hyper-parameter tuning was done by taking a small sub-sample of 50 OpenPose processed videos videos (25 ASD and 25 TD). We tested a ResNet50 CNN ([51]) for as the first CNN architechture for extraction of high dimensional features and a 512 LSTM unit recurrent neural network([45]) to add the temporal dimension for making video classification neural network. Out first test involved splitting the videos into segments of length 5, 10 and 15 seconds and monitored the training and validation loss to check for overfitting. We observed 5 second segments to provide us with the least validation loss among the 3 tests (see Fig.S2A). We further tested for improvements in the validation loss when the video dataset was downsampled to the resolution 320X240 from 696X554 resolution (see Fig.S2B) and observed a better fit for downsampled 320X240 videos. After conducting the previous tests we observed that a batch size of 256 or 128 was optimal to achieve the least validation loss. We also tested out several convolutional neural network architectures for feature extraction, excluding fully connected dense layers (see Fig.S2, panels C-D), out of which VGG16 ([43]) gave substantially better results compared to other CNN feature extraction methods such as ResNet50 ([51]). We also tried to use a bidirectional LSTM ([52]) in order to check for any further decrease in validation loss. However, using a bidirectional LSTM led to over-fitting in the neural network (see Fig.S2E).

Thus we observed the best model for our dataset to be a VGG16 LSTM at 128 batch size, 100 epochs and 320X240 resolution video with a validation loss of 0.301. We carry out the training with this configuration at 80-20 and 70-30 splits and observed a better performance with 80-20 split. Fig.S3. A detailed architecture of the VGG16 LSTM neural network can also be represented by Fig.S1.

**Figure S1.**
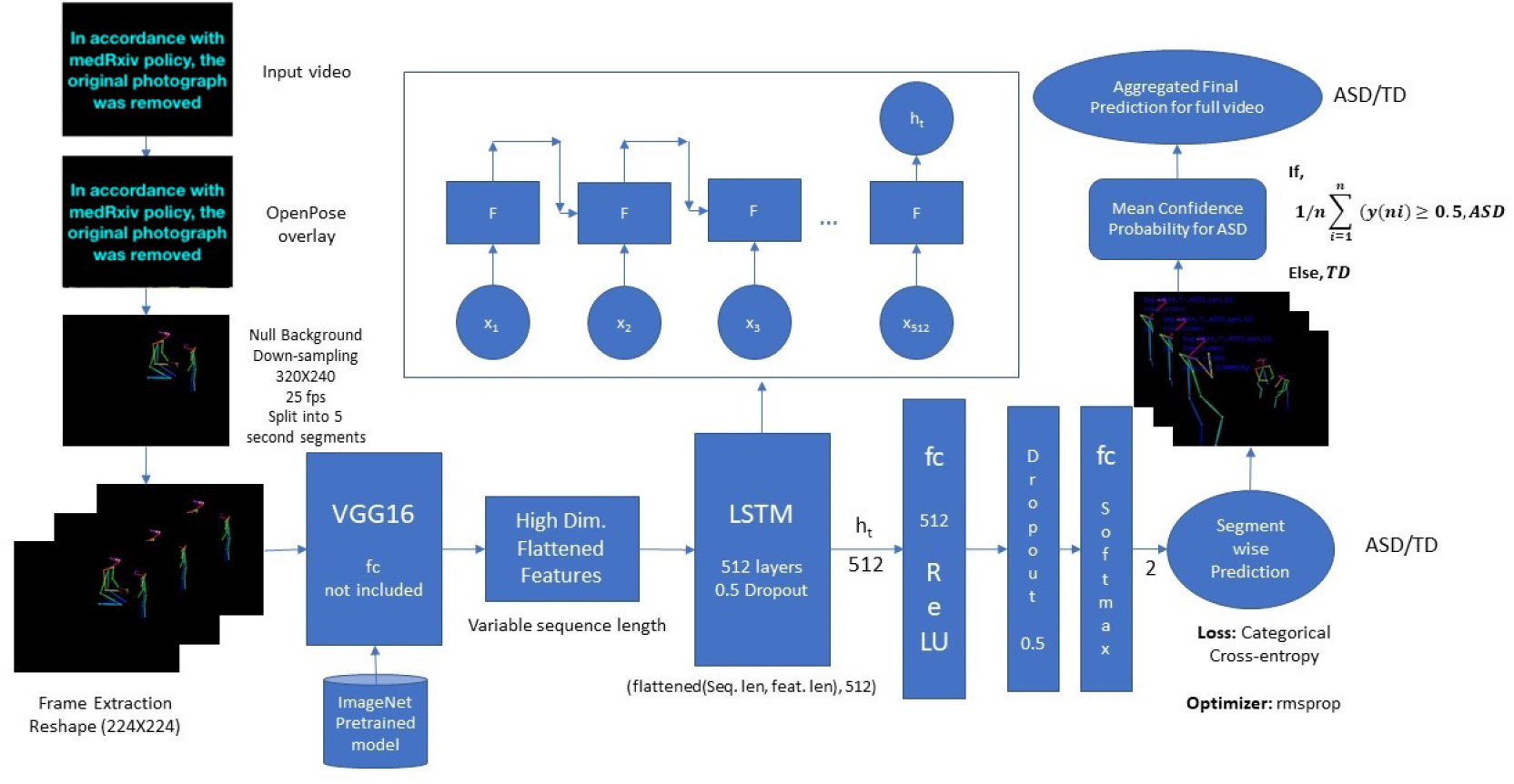
VGG16 LSTM detailed architecture representing pre-processing, feature extraction, addition of temporal dimension to features, segment-wise classification and aggregation of prediction over the entire video set to give final prediction.

**Figure S2.**
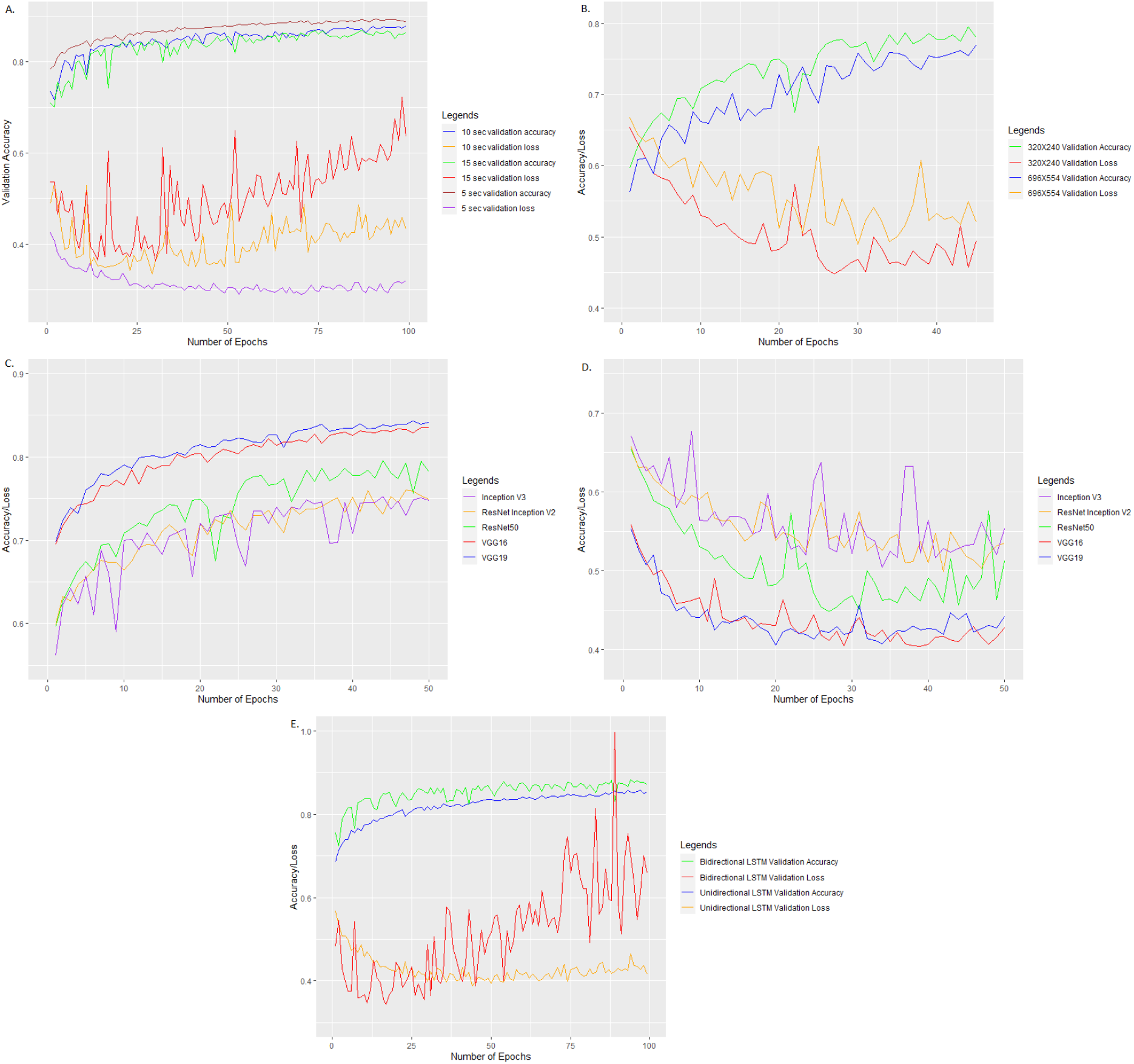
A. 5 sec, 10 sec and 15 second training and validation plots for ResNet50 LSTM at 256 batch size, 100 epochs and 70-30 training-validation split, B. 320X240 vs 696X554 resolution videos training and validation plots for ResNet50 LSTM, 256 batch size, 50 epochs, 70-30 training-validation split, C. Training and validation accuracy for different High Dimensional feature extraction CNN models with LSTM RNN model at 256 batch size, 50 epochs and 70-30 training-validation split, D. Training and validation accuracy for different High Dimensional feature extraction CNN models with LSTM RNN model at 256 batch size, 50 epochs and 70-30 training-validation split, E. Training and validation accuracy and loss for ResNet50 LSTM and ResNet50 Bidirectional LSTM at 100 epochs,256 batch size and 70-30 training-validation split

**Figure S3.**
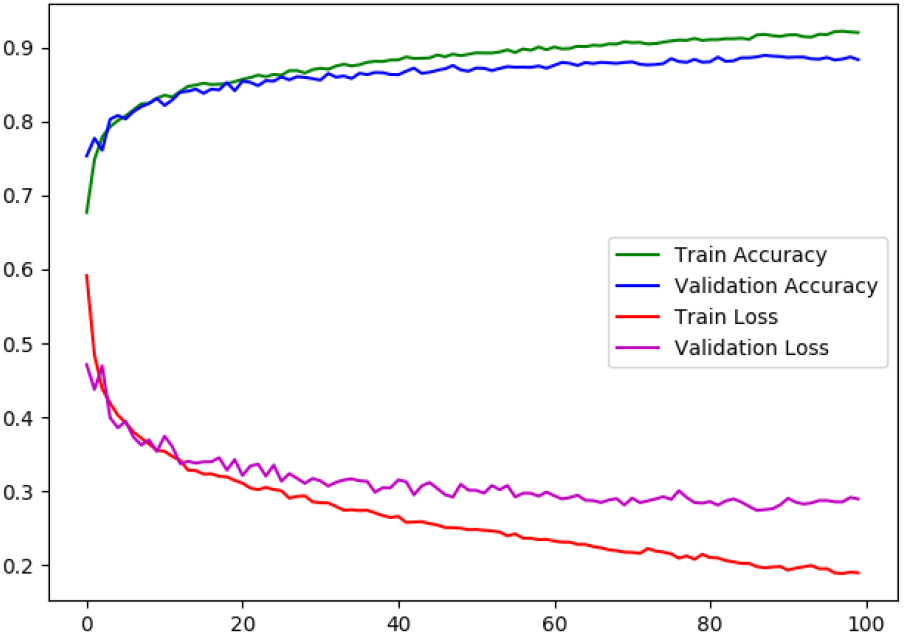
Training Accuracy, Training Loss, Validation Accuracy and Validation loss with 68 balanced training video dataset at 80-20 split, 128 Batch Size, 320X240 resolution and 100 Epochs for VGG16 LSTM neural network.

### Predicted class attribution across filming settings

The ADOS videos in the context of our longitudinal cohort were acquired using different systems. The majority of the videos was acquired using remotely controlled dome camera system. This setting involves cameras fixed at the height of 2 meters from the floor, and allowed manual pan, tilt, zoom as well as flexible switch between two camera views. This camera system was used in 44 videos from the training sample and 48 videos of the Testing set 1. The other settings included a fixed angle gopro camera that was positioned either high (above 160 cm of the floor) (Training: 10 videos, Testing:9 videos) either low (70-80cm from the floor) (Training: 14, Testing 11 videos). The accuracy of prediction across settings was the highest in a low positioned fixed angle gopro cameras, followed by the accuracy value of 81% in the setting involving high positioned manually controlled cameras. Finally, the poorest accuracy (77%) characterized the setting with a fixed cameras and high filming angle (see Fig.S4).

**Figure S4.**
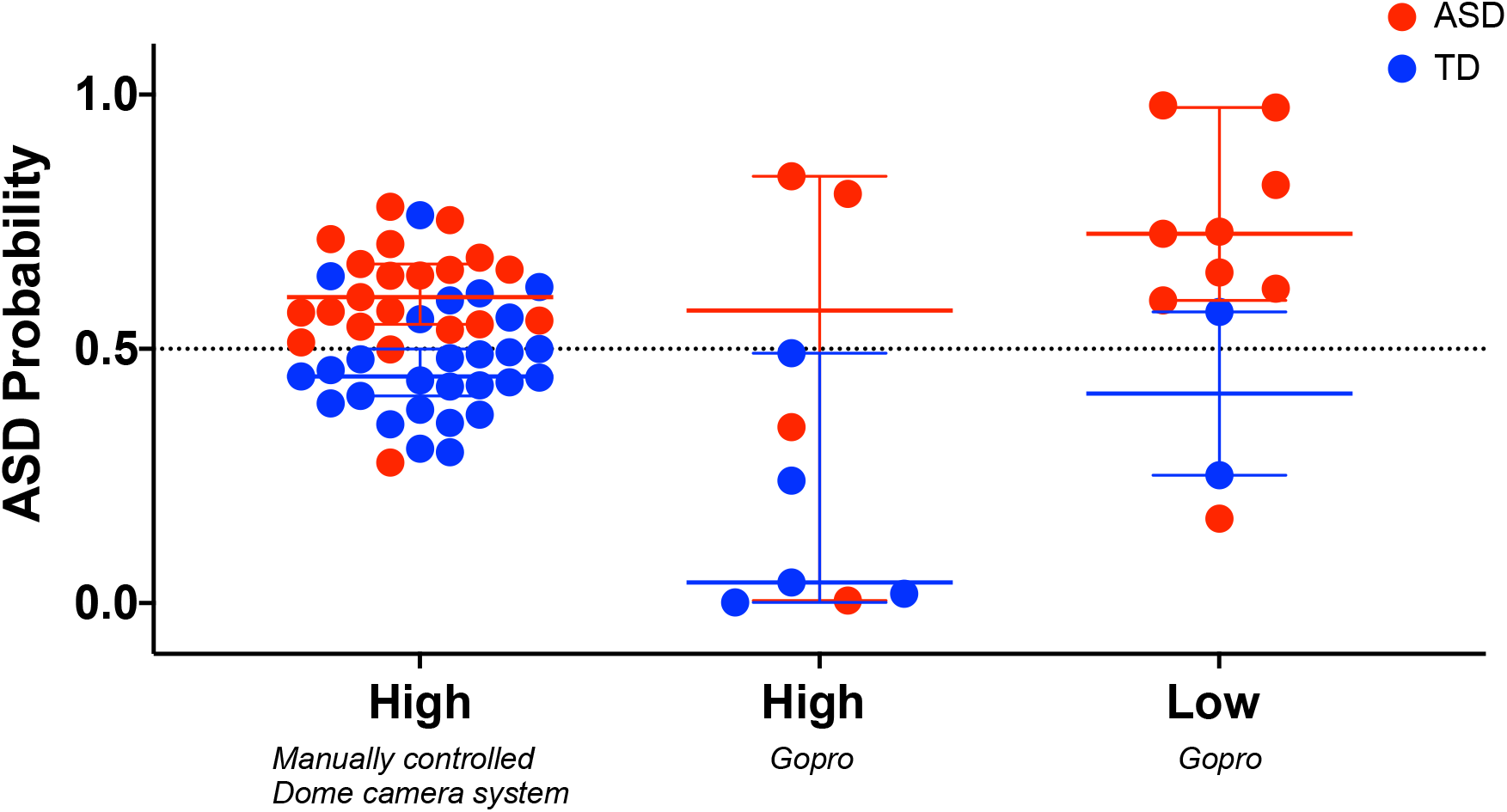
Scatter plots depicting ASD probability across the three setting types (columns) and for the two diagnostic groups (typically developing in blue and children with ASD in red). In high positioned manually controlled dome camera system the accuracy was 81%; in the setting involving gopros and high filming angle the accuracy was 77%; Finally, in the setting involving gopros positioned at low height filming angle the accuracy was 82%.

**Figure S5.**
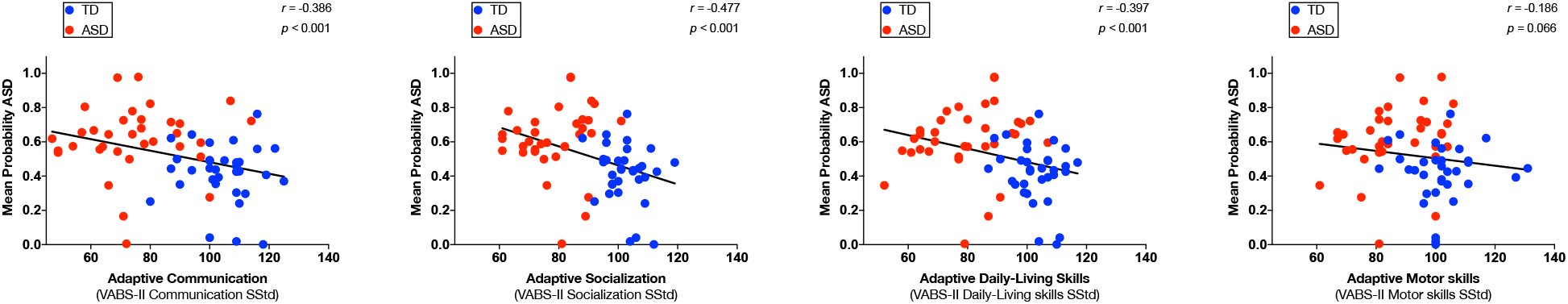
Relation between predicted ASD probability and subdomain of Vineland Adaptive Behavior Scales(p < 0.0125 after applying Bonferroni corrections for multiple comparisons). A. Communication domain; B. Socialization domain;C. Daily Living Skills domain; D. Motor domain. The least squares linear fit is depicted as black line and values of Spearman r coefficient and corresponding p values are shown on each graph.

**Figure S6.**
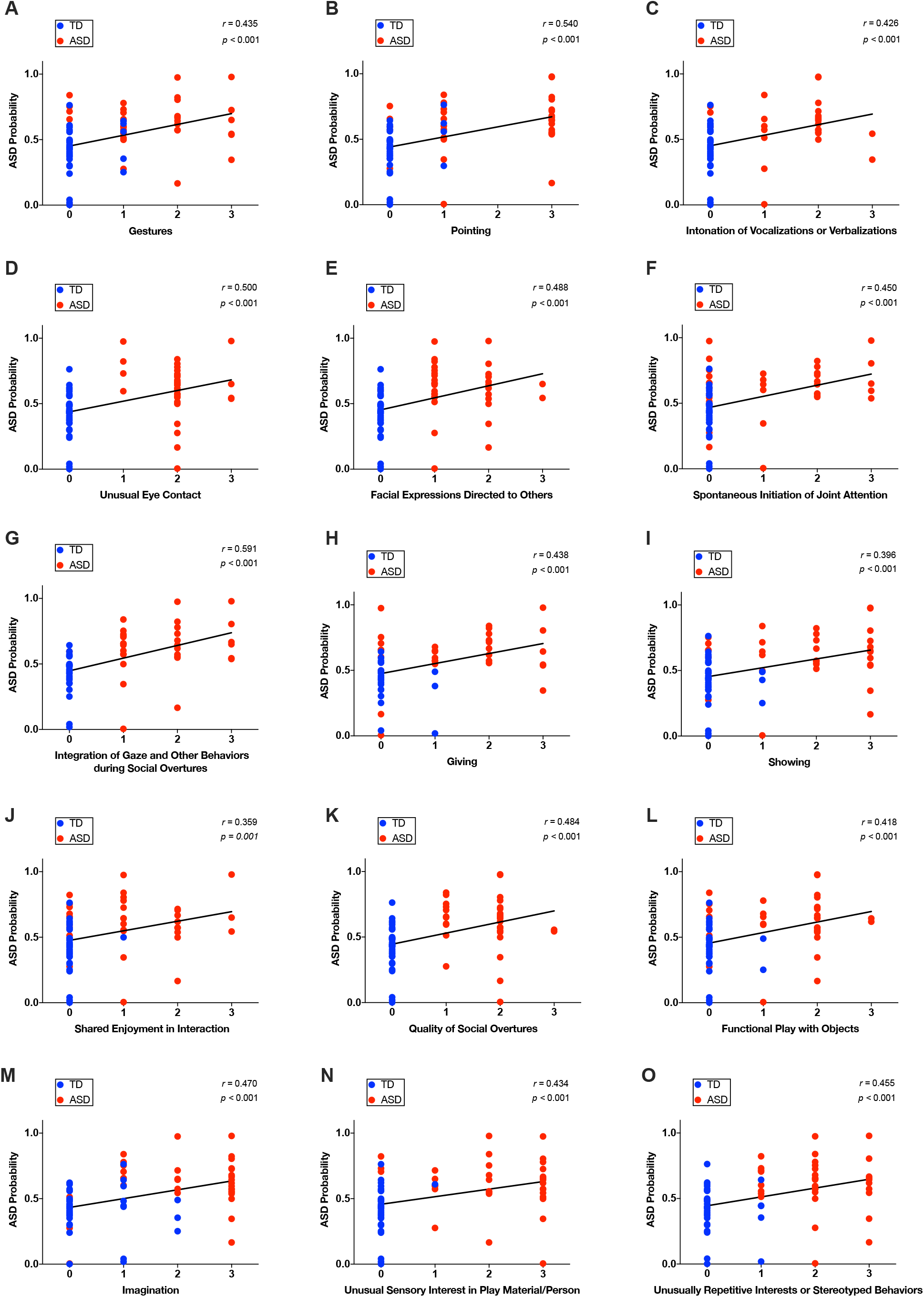
Relation between predicted ASD probability and individual symptoms derived from ADOS (p < 0.002 after applying Bonferroni corrections for multiple comparisons). A-C Communication domain; D-K Reciprocal Social Interaction domain; L-M Play; NO Repetitive and Restricted behaviors domain of ADOS. The least squares linear fit is depicted as black line and values of Spearman r coefficient and corresponding p values are shown on each graph.

In addition to testing the relation with general severity of symptoms of autism we also wanted to obtain the appreciation of the relation of ASD probability with the 27 individual symptoms that are coded across the three used ADOS modules (the list of symptoms included in this analysis is shown in Table S3). Of note, individual symptoms of autism in ADOS are scored on a 4 point scale ranging from 0 (“no evidence of abnormality”) to 3 (“markedly abnormal”).

**Table S2.** Selected items from the gold-standard diagnostic assessment, the ADOS-G [20] and ADOS-2 [21]. The different columns correspond to different modules the choice of which is performed as a function of age and language level.

| Behaviors | Toddler<br>Module | Module<br>1 | Module<br>2 |
| --- | --- | --- | --- |
| **A Communication** |  |  |  |
| Frequency of Spontaneous Vocalization Directed to Others | ☒ | ☒ | ☒* |
| Intonation of Vocalizations or Verbalizations | ☒ | ☒ | ☒ |
| Immediate Echolalia | ☒ | ☒ | ☒ |
| Stereotyped/Idiosyncratic Use of Words or Phrases | ☒ | ☒ | ☒ |
| Use of Another's Body | ☒ | ☒ | ☐ |
| Pointing | ☒ | ☒ | ☒ |
| Gestures | ☒ | ☒ | ☒ |
| **B Reciprocal Social Interaction** |  |  |  |
| Unusual Eye Contact | ☒ | ☒ | ☒ |
| Facial Expressions Directed to Others | ☒ | ☒ | ☒ |
| Integration of Gaze and Other Behaviors during Social Overtures | ☒ | ☒ | ☐ |
| Shared Enjoyment in Interaction | ☒ | ☒ | ☒ |
| Response to Name | ☒ | ☒ | ☒ |
| Requesting | ☒ | ☒ | ☐ |
| Showing | ☒ | ☒ | ☒ |
| Giving | ☒ | ☒ | ☐ |
| Spontaneous Initiation of Joint Attention | ☒ | ☒ | ☒ |
| Response to Joint Attention | ☒ | ☒ | ☒ |
| Quality of Social Overtures | ☒ | ☒ | ☒ |
| Amount of Social Overtures/Maintenance of Attention: Examiner | ☒ | ☒* | ☒* |
| Quality of Social Response | ☒ | ☒* | ☒ |
| Overall Quality of Rapport | ☒ | ☒* | ☒ |
| **C Play** |  |  |  |
| Functional Play with Objects | ☒ | ☒ | ☒ |
| Imagination / Creativity | ☒ | ☒ | ☒ |
| **D Stereotyped Behaviors and Restricted Interests** |  |  |  |
| Unusual Sensory Interest in Play Material/Person | ☒ | ☒ | ☒ |
| Hand and Finger and Other Complex Mannerisms | ☒ | ☒ | ☒ |
| Self-injurious behavior | ☒ | ☒ | ☒ |
| Unusually Repetitive Interests or Stereotyped Behaviors | ☒ | ☒ | ☒ |

\* Item not included in the given module of the older version (ADOS-G) of the schedule.

